# Perceptions, Attitudes, and Lived Experiences of Therapists with Lower Limb Robotic Exoskeletons in Stroke Rehabilitation: A Protocol for a Systematic Review of Qualitative Studies

**DOI:** 10.1101/2025.06.08.25329209

**Authors:** Ravi Shankar, Ziyu Goh, Xu Qian

## Abstract

Stroke is a leading cause of disability worldwide, often resulting in impaired walking ability. Lower limb robotic exoskeletons are an emerging technology in stroke rehabilitation that may enhance walking recovery. However, little is known about therapists’ perceptions, attitudes, and experiences with robotic exoskeletons. This protocol outlines a systematic review that aims to synthesize qualitative evidence on the perceptions, attitudes, and lived experiences of physical therapists and occupational therapists who use lower limb robotic exoskeletons in stroke rehabilitation.

The review will follow the Joanna Briggs Institute (JBI) methodology for systematic reviews of qualitative evidence. Eight electronic databases will be searched: PubMed, Web of Science, Embase, CINAHL, MEDLINE, The Cochrane Library, PsycINFO, and Scopus. Qualitative studies published in peer-reviewed journals that report on the perceptions, attitudes, or experiences of physical therapists and/or occupational therapists who have used lower limb robotic exoskeletons for stroke rehabilitation will be included. Two reviewers will independently screen studies, extract data, and assess methodological quality using the JBI Critical Appraisal Checklist for Qualitative Research. Thematic synthesis will be used to analyze and interpret the data.

Understanding therapists’ perceptions is crucial for the successful integration of robotic exoskeletons into clinical practice. Findings from this review will provide insights to guide implementation, training, and support for therapists using this technology. Results will inform an understanding of potential barriers and facilitators that can be used to optimize the use of robotic exoskeletons in stroke rehabilitation. This review will make a unique contribution by synthesizing qualitative evidence on an emerging topic with important implications for stroke rehabilitation practice.

## Introduction

Stroke is a leading cause of adult disability worldwide, with walking impairment being one of the most common post-stroke deficits [1]. Over 50% of stroke survivors experience long-term walking limitations [2], which can restrict participation in activities of daily living, community engagement, and quality of life [3]. Rehabilitation post-stroke aims to optimize functional recovery and independence. Conventional gait rehabilitation often involves intensive, repetitive task-specific training with manual assistance from one or more therapists [4]. However, this approach is physically demanding for therapists and difficult to sustain at the intensity required to drive neuroplastic changes [5].

Technological advances have led to the development of robotic devices for rehabilitation, including lower limb robotic exoskeletons [6]. Robotic exoskeletons are wearable, powered devices that strap around the user’s lower limbs to assist or enhance movement [7]. For stroke rehabilitation, robotic exoskeletons can provide intensive, high-dosage, repetitive gait training while reducing the physical burden on therapists [8]. A growing body of evidence supports the efficacy of robotic exoskeletons for improving walking speed, endurance, balance, and mobility in stroke survivors [9-11].

Despite the increasing use of robotic exoskeletons in research and clinical practice, little is known about therapists’ perceptions, attitudes, and experiences with this technology. Therapists play a crucial role in the clinical implementation of robotic exoskeletons, as they determine the suitability of patients, set up and apply the devices, oversee gait training, monitor patient safety and well-being, and assess outcomes [12]. Understanding therapists’ perspectives is essential for the successful integration of robotic exoskeletons into stroke rehabilitation practice.

Several studies have qualitatively explored therapists’ experiences with upper limb rehabilitation robots [13-15], but no reviews have synthesized the evidence on lower limb robotic exoskeletons. A systematic review of qualitative studies is warranted to gain a comprehensive understanding of therapists’ perceptions, as individual studies may be context-specific or have small sample sizes. Synthesizing qualitative evidence across different settings and devices can reveal overarching themes and novel insights to inform clinical practice, research, and technology development.

This protocol outlines a systematic review that aims to identify, appraise, and synthesize qualitative research evidence on the perceptions, attitudes, and lived experiences of physical therapists and occupational therapists who use lower limb robotic exoskeletons in stroke rehabilitation. The review will adhere to the JBI methodology for systematic reviews of qualitative evidence [16,17] and follow the Preferred Reporting Items for Systematic Reviews and Meta-Analyses (PRISMA) guideline [18]. The findings will make a unique contribution to the field by providing an in-depth understanding of therapists’ perspectives to guide the clinical implementation of robotic exoskeletons in stroke rehabilitation.

### Review question

The objective of this systematic review is to synthesize qualitative evidence that addresses the following question: What are physical therapists’ and occupational therapists’ perceptions, attitudes, and lived experiences of using lower limb robotic exoskeletons in stroke rehabilitation?

## Methods

The proposed systematic review will be conducted in accordance with the JBI methodology for systematic reviews of qualitative evidence [16,17].

### Inclusion criteria Participants

This review will consider studies that include licensed physical therapists and/or occupational therapists who have used a lower limb robotic exoskeleton for stroke rehabilitation, regardless of the specific device, setting, or level of experience. Studies focusing on other health professionals, such as rehabilitation assistants or engineers, will be excluded unless data from physical therapists and/or occupational therapists can be separately extracted.

### Phenomena of interest

The phenomena of interest for this review are therapists’ perceptions, attitudes, and lived experiences related to using lower limb robotic exoskeletons for gait rehabilitation after stroke. This encompasses a wide range of subjective perspectives and experiential insights related to the use of robotic exoskeletons in rehabilitation settings. It includes participants’ beliefs, opinions, understandings, and reflections on various aspects of the technology. For instance, these may involve perceived benefits, risks, barriers, and facilitators to use; the impact on clinical reasoning, treatment planning, and integration with conventional therapies; and how the technology influences therapist-patient interactions. Additionally, experiences related to device setup, programming, donning and doffing, as well as challenges faced and strategies employed to address them, are considered. Perspectives on training, confidence in using the exoskeleton, and the broader organizational, environmental, and attitudinal factors affecting adoption are also included. Finally, it covers therapists’ perceptions of patient experiences, clinical outcomes, and satisfaction with robotic exoskeleton-assisted interventions.

### Context

This review will consider studies conducted in any clinical setting where lower limb robotic exoskeletons are used for stroke rehabilitation, including inpatient rehabilitation, outpatient clinics, and community settings, without restrictions on geographical location.

### Types of studies

This review will consider primary qualitative studies that focus on the perceptions, attitudes, or lived experiences of therapists using lower limb robotic exoskeletons. These may include, but are not limited to, designs such as phenomenology, grounded theory, ethnography, action research, and qualitative descriptive studies. Mixed methods studies that include a distinct qualitatively-driven component will also be considered if the qualitative element addresses the phenomenon of interest. Studies published in peer-reviewed journals in any language will be included. Conference abstracts, opinion pieces, and commentaries will be excluded.

### Search strategy

The search strategy will aim to locate published studies from database inception to the present. An initial limited search of MEDLINE will be undertaken to identify relevant articles on the topic. The text words contained in the titles and abstracts of relevant articles, and the index terms used to describe the articles, will be used to develop a full search strategy. The search strategy, including all identified keywords and index terms, will be adapted for each included database. The reference lists of all included studies will be screened for additional papers.

The databases to be searched include: PubMed, Web of Science, Embase, CINAHL, MEDLINE, The Cochrane Library, PsycINFO, and Scopus.

A draft search string to be used is:

((“lower limb” OR “lower extremity” OR “leg” OR “robotic exoskeleton*” OR “powered exoskeleton*” OR “wearable robot*” OR “exoskeleton device*” OR “exoskeleton-assist*” OR “robot-assist*”) AND (“stroke” OR “cerebrovascular accident*” OR CVA OR “cerebrovascular disorder*”) AND (“physical therap*” OR physiotherap* OR “occupational therap*” OR rehabilitation) AND (perception* OR perspective* OR experience* OR attitude* OR view* OR opinion* OR reflection* OR belief* OR understandi ng*) AND (qualitative OR “mixed method*” OR interview* OR “focus group*” OR phenomenolog* OR “grounded theory” OR ethnograph* OR “action research” OR “qualitative descriptive” OR “thematic analysis” OR “content analysis” OR “narrative analysis” OR “discourse analysis”))

The search strategy will be broad in order to capture all relevant studies. No language or date restrictions will be applied. The searches will be re-run just before the final analyses to retrieve any additional recently published articles for inclusion.

### Study selection

Following the search, all identified citations will be collated and uploaded into Covidence (Veritas Health Innovation, Melbourne, Australia) and duplicates removed. Titles and abstracts will then be screened by two independent reviewers for assessment against the inclusion criteria for the review. Potentially relevant studies will be retrieved in full and their full text assessed in detail against the inclusion criteria by two independent reviewers. Any disagreements between the reviewers at each stage of the selection process will be resolved through discussion, or with a third reviewer. Reasons for exclusion of full-text studies that do not meet the inclusion criteria will be recorded and reported in the systematic review. The results of the search will be reported in full in the final systematic review and presented in a PRISMA flow diagram [19].

### Assessment of methodological quality

Eligible studies will be critically appraised by two independent reviewers for methodological quality using the standard JBI Critical Appraisal Checklist for Qualitative Research [16]. The checklist used in this review evaluates the methodological rigor and coherence of qualitative studies across ten key criteria. These include assessing the alignment between the stated philosophical perspective and the chosen methodology, as well as consistency between the methodology and the research questions, data collection methods, data analysis, and interpretation of results. It also examines whether the researcher is positioned culturally or theoretically, and whether their influence on the research—and vice versa—is acknowledged. The checklist further evaluates the extent to which participants and their voices are meaningfully represented, the presence of ethical considerations and approval, and whether the study’s conclusions logically follow from the data analysis.

Studies will be assessed as having met, not met, or unclear for each individual criterion. Those meeting all 10 criteria will be considered of high methodological quality. Studies meeting 7 to 9 criteria will be deemed of moderate quality, while those meeting 6 or fewer criteria will be considered of low quality. Any disagreements that arise between the reviewers will be resolved through discussion or with a third reviewer. The appraisal results will be reported in narrative form and in a table, along with a description of the methodological quality of the included studies.

Importantly, studies will not be excluded based on methodological quality, as the aim is for an inclusive synthesis of the full range of available evidence. However, the quality appraisal results will be transparently reported and considered during data synthesis and interpretation of findings.

### Data extraction

Data will be extracted from included studies by two independent reviewers using a data extraction tool developed by the reviewers. The data extracted will include specific details about the study populations, context, culture, geographical location, study methods, phenomena of interest, outcomes, and findings relevant to the review objective. Authors of papers will be contacted to request missing or additional data where required.

The data extraction tool will be piloted on a small number of included studies and modified as needed before use on the remaining studies. Modifications will be detailed in the final systematic review report. Disagreements between the reviewers will be resolved through discussion or with a third reviewer.

### Data synthesis

Qualitative research findings will be pooled using thematic synthesis, as described by Thomas and Harden [20]. This involves three stages: 1) line-by-line coding of the findings of primary studies; 2) development of descriptive themes; and 3) generation of analytical themes.

In the first stage, the findings of each included study will be entered verbatim into NVivo qualitative data analysis software (QSR International Pty Ltd. Version 12, 2018). Data will be coded line-by-line to develop a bank of codes that describe the meaning and content of each extract. Codes will be inductively derived based on the data rather than using a pre-defined framework.

In the second stage, the codes will be examined for similarities and differences, and grouped into a hierarchical tree structure to generate descriptive themes and sub-themes. The tree structure will be refined through constant comparison and discussion within the review team until the synthesis captures the full range and depth of insights across the included studies.

In the third stage, higher-order analytical themes will be developed by examining patterns, relationships, and interpretations across the descriptive themes. The aim is to ‘go beyond’ a summary of the primary studies to generate new understandings that can directly inform clinical practice and policy. The review team will discuss the analytical themes to ensure they are grounded in the data but extend the original findings to produce a new interpretation.

Throughout the data synthesis process, the review team will meet regularly to discuss the emerging themes and interpretations. A reflexive approach will be taken, with reviewers considering how their own backgrounds, assumptions, and beliefs may influence the analysis. Detailed records will be kept to ensure transparency and enable auditing of the synthesis process.

### Rigor

Several strategies will be employed to enhance the trustworthiness and credibility of the data synthesis [21]. Firstly, a comprehensive search strategy will aim to locate all available relevant evidence. Secondly, appraisal of methodological quality will be used to identify potential limitations of the included studies. Thirdly, data extraction and coding will be conducted by two independent reviewers to reduce potential bias.

Fourthly, a detailed codebook will be developed and refined to ensure consistency in coding across the included studies. Fifthly, regular team meetings will provide a forum for critical discussion of the analysis and interpretation. Finally, the review findings will include rich descriptions and direct quotations from participants to ensure the synthesis remains grounded in the data.

## Discussion

This systematic review will synthesize qualitative research evidence on physical therapists’ and occupational therapists’ perceptions, attitudes, and experiences of using lower limb robotic exoskeletons in stroke rehabilitation. To our knowledge, this will be the first meta-synthesis to focus specifically on therapist perspectives of robotic exoskeletons.

The findings will contribute to a comprehensive understanding of therapist experiences across diverse practice settings and robotic devices. Synthesizing qualitative evidence can identify broader themes and patterns that may not be apparent in individual studies. This is particularly important for an emerging technology like robotic exoskeletons, where therapist perspectives may evolve as devices become more widely adopted in clinical practice.

Several potential implications and applications of the review findings can be envisioned. Firstly, the results may inform the development and refinement of clinical training programs to better prepare therapists to effectively integrate robotic exoskeletons into stroke rehabilitation. Understanding therapists’ learning needs, preferred training formats, and common challenges can help optimize educational initiatives. Secondly, the findings may guide device manufacturers to incorporate therapist-desired features or interfaces that enhance the clinical utility and usability of robotic exoskeletons. Thirdly, the review may identify organizational or health system factors that influence therapist uptake of robotic exoskeletons, informing targeted strategies to address barriers and leverage facilitators.

Additionally, the review may uncover important insights into the impact of robotic exoskeletons on therapist-patient interactions and relationships. While quantitative studies have demonstrated the efficacy of these devices for improving gait outcomes, less is known about potential shifts in rehabilitation dynamics and processes. Therapist reflections can highlight how their roles and therapeutic approaches may evolve when working with robotic technologies.

The proposed review has several methodological strengths, including adherence to best practice guidelines for the conduct and reporting of qualitative systematic reviews, a comprehensive search strategy encompassing published and gray literature, rigorous quality appraisal, and the use of established techniques for qualitative evidence synthesis. Nonetheless, some limitations should be acknowledged. The findings will be constrained by the quality of the included primary studies. The search will be limited to English language publications, potentially excluding valuable insights from other linguistic and cultural contexts. Additionally, the synthesis findings will need to be interpreted in light of the inherent subjectivity of qualitative research.

Future research could build upon this review in several directions. Conducting primary qualitative studies in regions or settings not represented in the current evidence base could enhance the transferability of the findings. Reviews focusing on the perspectives of other stakeholders, such as patients, caregivers, and rehabilitation managers, could allow for triangulation of experiences. Finally, developing and evaluating knowledge translation interventions based on the review findings could support the optimal implementation of robotic exoskeletons in clinical practice.

## Conclusion

This systematic review will provide timely and valuable insights into physical therapists’ and occupational therapists’ perceptions, attitudes, and experiences of using lower limb robotic exoskeletons in stroke rehabilitation. The findings will inform the development of clinical training, device design, and organizational supports to facilitate the successful adoption of this promising technology. By understanding therapist perspectives, we can develop targeted strategies to ensure robotic exoskeletons are effectively integrated into rehabilitation practice to optimize patient outcomes and experiences. Ultimately, the review aims to advance an evidence-based approach to the clinical implementation of robotic exoskeletons, grounded in the real-world knowledge of frontline therapists.

## Data Availability

All data produced in the present work are contained in the manuscript.

